## Supplementary for "Clinical characteristics and factors associated with COVID-19-related mortality and hospital admission in 5 rural provinces in Indonesia: a retrospective cohort study"

**Supplementary data**

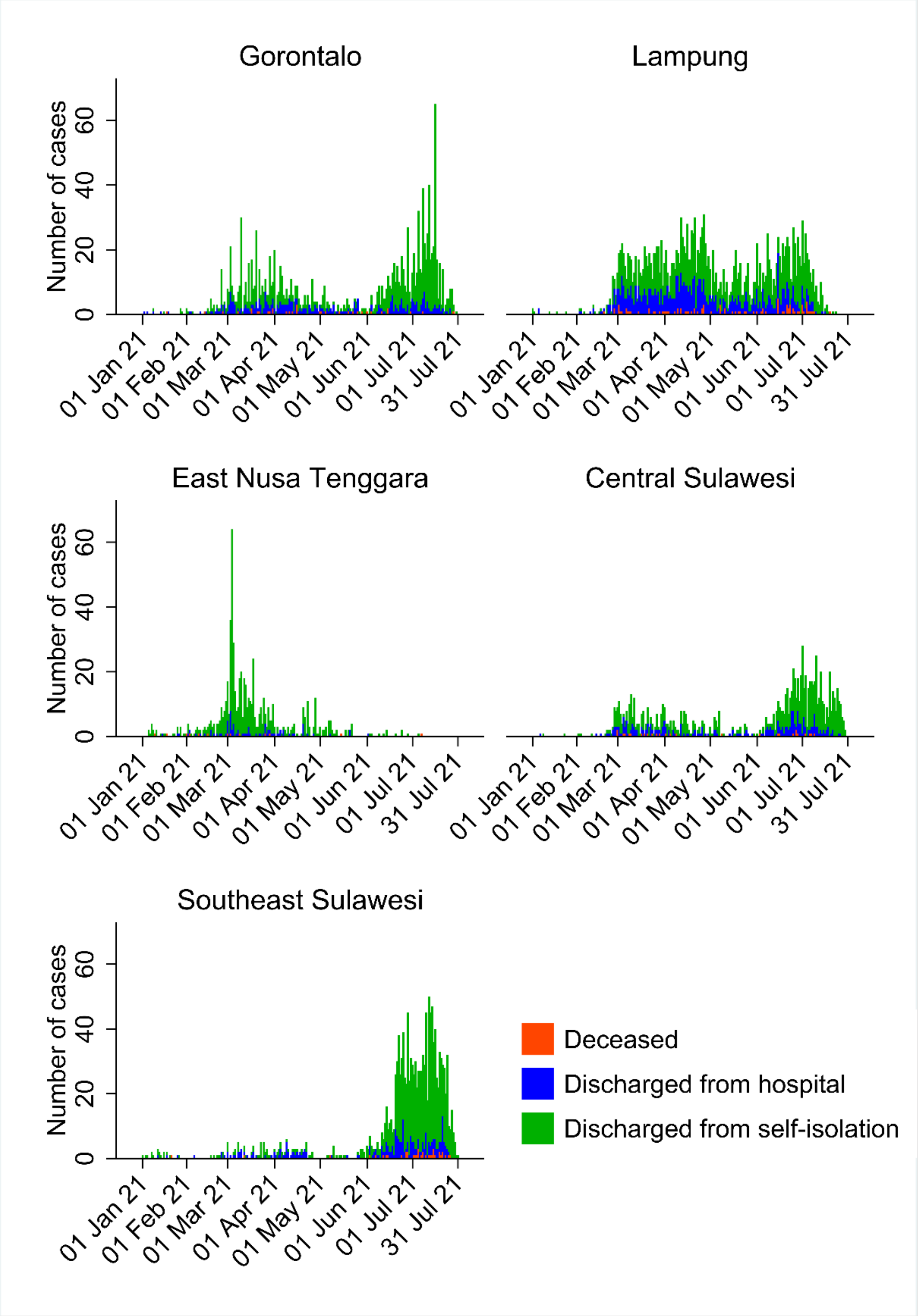


**Supplementary Figure 1: Number of COVID-19 outcomes over time by province**

**Supplementary Table 1. Population number and number of and health care workers by province**

| Province | Population number | Number of doctors per 100,000 population | Number of nurses per 100,000 population | Number of midwives per 100,000 population | Number of public health officer per 100,000 population |
|---|---|---|---|---|---|
| Lampung | 9,007,848 | 4.97 | 10.52 | 19.18 | 9.20 |
| Gorontalo | 1,171,681 | 6.57 | 17.41 | 9.13 | 51.40 |
| Central Sulawesi | 2,985,734 | 8.47 | 27.46 | 13.73 | 47.60 |
| Southeast Sulawesi | 2,624,875 | 12.59 | 36.51 | 13.03 | 52.50 |
| East Nusa Tenggara | 5,325,566 | 6.80 | 25.63 | 12.74 | 14.00 |
| Jakarta | 10,562,088 | 44.81 | 48.30 | 30.70 | NA |

NA=Not available

**Supplementary Table 2: Bivariable mixed effects logistic regression analysis showing factors associated with risk of COVID-19 mortality and hospitalisation in five rural provinces, Indonesia**

| | **Mortality** | | **Hospitalisation** | |
|---|---|---|---|---|
| | **OR (95% CI)** | **p value** | **OR (95% CI)** | **p value** |
| **Demographics** | | | | |
| Age group, years | | | | |
| 0-19 | 1 (reference) | | 1 (reference) | |
| 20-29 | 2.20 (0.61-7.89) | 0.228 | 1·13 (0·88-1·44) | 0.340 |
| 30-39 | 2.40 (0.67-8.51) | 0.177 | **1·31 (1·03-1·67)** | **0.029** |
| 40-49 | **5.87 (1.76-19.61)** | **0.004** | **2·02 (1·59-2·57)** | **<0.0001** |
| 50-59 | **15.44 (4.81-49.51)** | **<0.0001** | **4·31 (3·41-5·45)** | **<0.0001** |
| 60-69 | **32.85 (10.24-105.34)** | **<0.0001** | **6·25 (4·83-8·10)** | **<0.0001** |
| ≥70 | **51.65 (15.74-169.51)** | **<0.0001** | **7·52 (5·41-10·47)** | **<0.0001** |
| Sex | | | | |
| Female | 1 (reference) | | 1 (reference) | |
| Male | **1.38 (1.05-1.83)** | **0.023** | 0·97 (0·87-1·09) | 0.623 |
| **Clinical characteristics** | | | | |
| Clinical diagnosis with pneumonia | **16.13 (11.51-22.62)** | **<0.0001** | **22·50 (16·01-31·62)** | **<0.0001** |
| Type of comorbidity | | | | |
| Hypertension | **6.14 (4.52-8.34)** | **<0.0001** | **5·42 (4·49-6·54)** | **<0.0001** |
| Diabetes | **7.80 (5.63-10.81)** | **<0.0001** | **6·44 (5·11-8·11)** | **<0.0001** |
| Cardiac diseases | **7.16 (4.72-10.85)** | **<0.0001** | **5·96 (4·35-8·15)** | **<0.0001** |
| COPD | | | **15·04 (6·67-33·90)** | **<0.0001** |
| Chronic kidney diseases | **8.90 (4.45-17.83)** | **<0.0001** | **9·34 (4·57-19·11)** | **<0.0001** |
| Liver diseases | **12.91 (3.98-41.90)** | **<0.0001** | **11·40 (3·10-41·89)** | **<0.0001** |
| Malignancy | **7.66 (2.52-23.29)** | **<0.0001** | **3·13 (1·27-7·69)** | **0.013** |
| Immunocompromised | NA | NA | **5·26 (1·92-14·42)** | **<0.0001** |
| Number of comorbidities | | | | |
| 0 | 1 (reference) | | 1 (reference) | |
| 1 | **7.48 (5.37-10.41)** | **<0.0001** | **6·29 (5·32-7·44)** | **<0.0001** |
| >1 | **19.74 (13.55-28.75)** | **<0.0001** | **10·25 (7·62-13·78)** | **<0.0001** |

| | | | | |
|---|---|---|---|---|
| **Health care workers** | | | | |
| Number of doctors per 100,000 population | 0.63 (0.28-1.39) | 0.250 | 0.47 (0.13-1.71) | 0.254 |
| Number of nurses per 100,000 population | **0.82 (0.68-0.99)** | **0.037** | **0.72 (0.53-0.97)** | **0.029** |
| Number of midwives per 100,000 population | **2.00 (1.38-2.91)** | **<0.0001** | 2.07 (0.78-5.48) | 0.144 |
| Number of public health officers per 100,000 population | 0.91 (0.83-1.01) | 0.068 | 0.98 (0.80-1.19) | 0.822 |

Province was treated as the random effect variable.
OR: odds ratio.
NA: Not Applicable due to 0 value of observation with immunocompromised condition among deceased cases.

**Supplementary Table 3: Mixed effects logistic regression multivariable models assessing association between and mortality and hospitalisation with number of comorbidities in five rural provinces, Indonesia**

| | **Mortality** | | **Hospitalisation** | |
|---|---|---|---|---|
| | **aOR (95% CI)** | **p value** | **aOR (95% CI)** | **p value** |
| Number of comorbidities | | | | |
| 0 | 1 (reference) | | 1 (reference) | |
| 1 | **2.56 (1.68-3.90)** | **<0.0001** | **3·56 (2·91-4·35)** | **<0.0001** |
| >1 | **4.95 (3.01-8.15)** | **<0.0001** | **4·46 (3·14-6·35)** | **<0.0001** |

Model was adjusted for age, sex, pneumonia, and number of health care workers per 100,000 population. Province was treated as the random effect variable. aOR: adjusted odds ratio.
